# PSYCHOLOGICAL READINESS FOLLOWING ANTERIOR CRUCIATE LIGAMENT INJURY AND REINJURY IN ADOLESCENTS AND YOUNG ADULTS: A RETROSPECTIVE COHORT STUDY IN SPORTS PHYSICAL THERAPY CLINICS

**DOI:** 10.64898/2026.03.06.26347203

**Authors:** Jordan D. Moser, Colin W. Bond, Benjamin C. Noonan

## Abstract

**Objectives:** Compare Anterior Cruciate Ligament (ACL) Return to Sport after Injury (ACL-RSI) scores over time following ACL reconstruction (ACLR) between male and female patients aged 15 to 25 years with primary ACL injuries and ACL reinjuries.

**Design:** Retrospective cohort design.

**Setting:** Sports physical therapy clinics.

**Participants:** 332 patients aged 15-25 years who underwent ACLR following either primary ACL injury or ACL reinjury, either contralateral or ipsilateral graft reinjury, and had at least one observation of the ACL-RSI.

**Main Outcome Measures:** ACL-RSI score.

**Results:** ACL-RSI scores significantly increased over time post- ACLR (p < .001), males reported significantly higher scores compared to females (p < .001), and patients with contralateral ACL reinjury demonstrated higher scores than those with ipsilateral ACL graft reinjury (p = .006), though there was no difference in scores between patients with primary ACL injury and ACL reinjury. A significant interaction effect of sex and injury status was also observed (p = .009), generally demonstrating that females had lower psychological readiness compared to males across injury statuses.

**Conclusions:** ACL-RSI following ACLR varies based on biological sex and time post-ACLR, though ACL reinjury, independent of the reinjured leg, does not appear to effect scores compared to primary ACL injury.

## INTRODUCTION

A patient’s psychological response to anterior cruciate ligament (ACL) injury and psychological recovery after ACL reconstruction (ACLR) can impact outcomes such as return to sport (RTS). It is not well understood how this response and recovery differs between patients following primary ACL injury, contralateral ACL reinjury, and ipsilateral ACL graft reinjury.(Ashton et al., 2020; Duncan et al., 2023; Momaya et al., 2024; Obradovic et al., 2024) Improving this understanding could enhance the ability to characterize a patient’s psychological readiness and potentially improve outcomes.

Patient’s may exhibit a number of maladaptive psychological responses to ACL injury, which include but is not limited to depression, anxiety, fear of reinjury, a loss of self-efficacy, and avoidance behaviors.(Ashton et al., 2020; Burland et al., 2019; Faleide & Inderhaug, 2023; Faleide et al., 2021; Hsu et al., 2017; Milewski et al., 2023; Paterno et al., 2018; Webster & Feller, 2022; Webster et al., 2018) These responses may be caused by a strong athletic identity and the associated disruption to that identity, pain, perceived lack of control of the situation or external locus of control, unrealistic expectations, poor social support, a patient’s individual emotional makeup and coping style, hyperawareness of the injured joint, or exacerbation of a pre-existing condition among other factors. (Ashton et al., 2020; Burland et al., 2019; Faleide & Inderhaug, 2023; Faleide et al., 2021; Hsu et al., 2017; Milewski et al., 2023; Paterno et al., 2018; Webster & Feller, 2022; Webster et al., 2018)

For patients aiming to RTS, psychological readiness is a critical factor that must be measured and addressed as part of the rehabilitation and decision-making process.(Faleide et al., 2021) The Anterior Cruciate Ligament – Return to Sport after Injury (ACL-RSI) is one tool that is used frequently to assess a patient’s psychological readiness to RTS, encompassing three domains: emotions, confidence in performance, and risk appraisal.(Ashton et al., 2020; Barth et al., 2023; Burland et al., 2019; Faleide & Inderhaug, 2023; Faleide et al., 2021; Hsu et al., 2017; Obradovic et al., 2024; Sell et al., 2024; Webster et al., 2008; Xiao et al., 2023; Zwolski et al., 2023) The 12-item scale asks participants to rate themselves on a continuum from "not at all" to "extremely".(Webster et al., 2008) Although the ACL-RSI measures three psychological domains, it is considered unidimensional, and an overall score is calculated by averaging the 12 items, with scores ranging from 0 to 100%, where higher scores are suggested to indicate greater psychological readiness.(Webster et al., 2008) The ACL-RSI scale is considered valid and reliable for use with both adolescent and adult populations.(Cirrincione et al., 2023)

For patients following primary ACL injury, ACL-RSI scores gradually increase with time reaching a plateau phase between 6 and 24 months post-ACLR with some patients experiencing a “second wave” of maladaptive psychological response just before they are ‘cleared’ to RTS.(Barth et al., 2023; Sell et al., 2024; Webster et al., 2008; Webster et al., 2018; Xiao et al., 2023) Patients who sustain an ACL reinjury, either to their contralateral native ACL or ipsilateral ACL graft, may exhibit even lower ACL-RSI scores after the second injury. Duncan et al. (2023) demonstrated that ACL revision patients exhibit an ACL-RSI score approximately 8% lower at the time of RTS compared to patients after primary ACLR. This difference may stem from several factors, including heightened fear of reinjury—specifically the fear of a third ACL injury—given their reality of having already experienced reinjury, reduced optimism about their second recovery process, negative comparisons between their first and current rehabilitation, further diminished confidence, the daunting challenge of repeating rehabilitation and its impact on motivation, and unresolved maladaptive responses to the initial injury, all of which contribute to the disruption of their athletic identity due to the additional time lost.(Duncan et al., 2023) These factors are particularly plausible given the proposed link between low ACL-RSI scores following primary ACL injury and an increased risk of reinjury.(Duncan et al., 2023; McPherson et al., 2019a, 2019b)

Further research is needed to understand the differences in ACL-RSI scores and its trajectory over time post-ACLR between patients with primary ACL injuries and those with reinjuries, as well as how these differences are influenced by contralateral ACL reinjury or ipsilateral ACL graft reinjury.(Duncan et al., 2023) Further, for studies assessing psychological readiness after primary ACL injury, many report ACL-RSI scores from samples with a mean age much older than the high-risk population typically seeking a return to cutting and pivoting sports, only assess ACL-RSI at RTS rather than its trajectory over time post-ACLR, and do not compare male and female patients when previous research has identified that females exhibit lower ACL-RSI scores than males.(Barth et al., 2023; Bruder et al., 2023; Duncan et al., 2023; Kostyun et al., 2021; Milewski et al., 2023; Obradovic et al., 2024; Sell et al., 2024; Xiao et al., 2023)

Accordingly, the purpose of this study is to compare ACL-RSI scores over time post-ACLR in male and female patients 15-25 years of age after primary ACL injury and ACL reinjury, to either the contralateral native ACL or ipsilateral ACL graft. We hypothesized that ACL-RSI score will be lower following the ACL reinjury compared to the primary ACL injury, with females exhibiting lower scores than males after both the primary ACL injury and ACL reinjury.

## METHODS

### Subjects

The (removed to remain anonymous for peer review) Institutional Review Board granted the retrospective analysis an exemption (review number: 2390). Beginning in 2012, post-ACLR data from patients who completed return to play (RTP) tests at our health system’s sports physical therapy clinics were added to our database. Anonymous data was provided by an honest broker on January 23^rd^, 2025. At this time, the full database contained data from 1,125 ACLR patients with a combined 2,019 RTP tests representing 63 surgeons. Patients who underwent ACLR for primary ACL injury, subsequently sustained a contralateral ACL or ipsilateral ACL graft reinjury and underwent ACLR again were included in this analysis if they had at least one observation of ACL-RSI after the first or second ACL injury. Patients were included regardless of articular cartilage or menisci pathologies treated at the time of ACLR. Patients were excluded from the analysis if they were younger than 15 or older than 25 years at the time of ACLR, had an allograft ACLR, or had the autograft harvested from the non-ACL injured leg for the primary ACLR. After exclusion, 332 ACLR patients with a combined 561 post-ACLR RTP tests were included in this retrospective analysis.

### Procedures

From 2012 to 2024, our healthcare system performed approximately 500 ACLR cases annually. Many of these patients are referred to our health care system’s sports physical therapy facilities for rehabilitation and/or one or more post-ACLR RTP tests. Additionally, patients who had their ACLR performed at other health care facilities may be referred to our facilities for physical rehabilitation and post-ACLR RTP tests. Patients may complete one or more post-ACLR RTP tests, typically between 3 and 9 months post-ACLR. Components of the post-ACLR RTP test include a variety of patient reported outcome measures, including but not limited to the ACL-RSI.

The 12 question ACL-RSI was completed. Patients responded to the ACL-RSI questions by circling a number on a 0 to 100-point scale provided in 10-point increments using prompts associated with each end of the scale’s continuum. The ACL-RSI is considered unidimensional and the mean score, presented as proportional data on a scale from 0% (maladaptive emotions, performance confidence, and risk appraisal) to 100% (adaptive emotions, performance confidence, and risk appraisal), for its 12 questions was subsequently computed. Patients initially completed the ACL-RSI on paper and the data was subsequently transferred to a digital database managed using REDCap.

### Statistical Analysis

The continuous dependent variable of this study was ACL-RSI score. The continuous independent variable of this study was time post-ACLR, measured in months, and the categorical independent variables of this study were biological sex and injury status, which was either primary injury or reinjury, with reinjury subcategorized as contralateral reinjury or ipsilateral graft reinjury. Prior to model fitting, the ACL-RSI score was transformed from the probability scale to the logit scale. We employed a linear mixed-effects model to analyze the influence of time post-ACLR, biological sex, and injury status as well as their interaction terms on logit transformed ACL-RSI scores. The model incorporated a random effect for patient to account for inter-patient variability, acknowledging that some patients in the data set had observations after primary ACL injury and ACL reinjury and repeated measures from the same patient are potentially correlated. To model the covariance structure of the repeated measures, we specified a continuous autoregressive correlation structure, which assumes that measurements taken closer in time are more highly correlated than those taken further apart. We also addressed potential heteroscedasticity by applying a combination of variance functions allowing for different variances across levels biological sex and injury status. The distribution of the model’s standardized residuals was robustly assessed to confirm normality. Analysis of variance was used to assess the effect of each independent variable and their interaction terms on logit transformed ACL-RSI scores. Bonferroni adjusted post-hoc comparisons were then computed on the logit scale where appropriate and the estimated marginal means were reverse transformed to the probability scale for illustration. Significance was set at p < 0.05. Statistical analysis was completed in R (v. 4.2.2). Based on the results of Duncan et al. (2023) who reported an ACL-RSI score of 85.3% ± 17.4 for primary ACLR patients and 77.4% ± 19.4 for revision ACLR patients at the time of return to sport with significance set at p < 0.05 and a desired power of 0.80, 87 primary ACLR patients and 87 reinjury ACLR patients were required.

## RESULTS

Demographic data are presented in Table 1. The ANOVA revealed an effect of time post-ACLR (F = 97.1, p < .001), biological sex (F = 22.9, p < .001), injury status (F = 7.4, p < .001), and a biological sex by injury status interaction (F = 4.7, p = .009). ACL-RSI scores increased with time post-ACLR and males had higher ACL-RSI scores compared to females. Patients with a contralateral ACL reinjury had higher ACL-RSI scores than patients with an ipsilateral ACL graft reinjury (p = .006), but there was no difference in ACL-RSI scores between patients with a primary ACL injury and either a contralateral (p = .069) or ipsilateral ACL graft reinjury (p = .179). Post-hoc test results for the biological sex by injury status interaction are presented in Table 2. Estimated marginal means as a function of time post-ACLR for male and female primary ACL injury, contralateral ACL reinjury, and ipsilateral ACL graft reinjury patients are provided in Figure 1.

**Figure 1.**
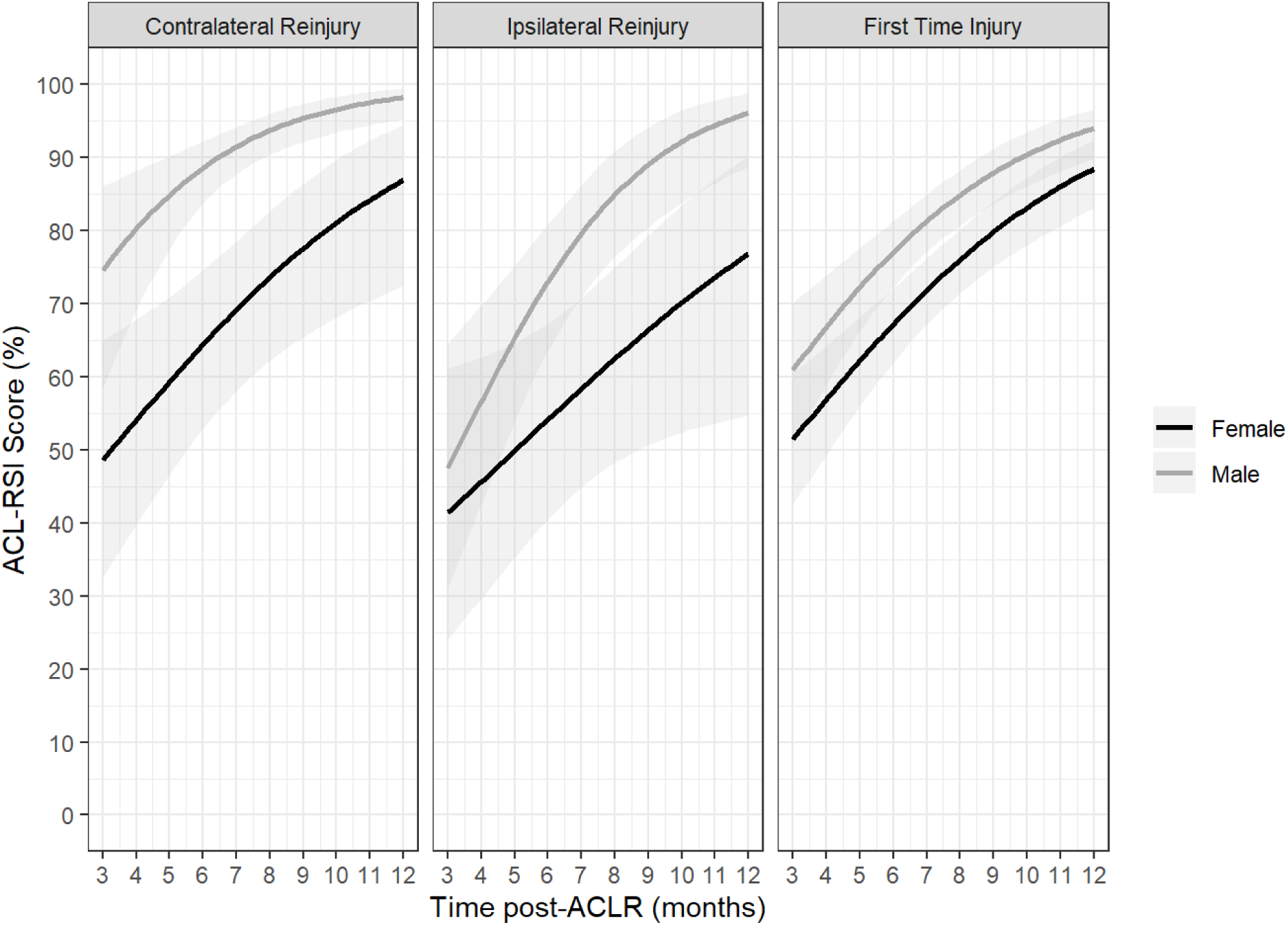
Estimated marginal means as a function of time post-anterior cruciate ligament (ACL) reconstruction for male (grey) and female (black) primary ACL injury, contralateral ACL reinjury, and ipsilateral ACL graft reinjury patients. Solid line: mean; ribbon: 95% confidence interval for the mean.

**Table 1.** Demographic data.

| Primary ACL Injury<br>(n = 251) | Contralateral ACL Reinjury<br>(n = 42) | Ipsilateral ACL Graft Reinjury<br>(n = 53) |
| --- | --- | --- |
| **Sex (n)** |  |  |
| Male = 130 | Male = 26 | Male = 36 |
| Female = 121 | Female = 16 | Female = 17 |
| **Pre-Injury Marx (out of 16)** |  |  |
| 14.5 ± 2.8 | 14.5 ± 2.6 | 13.2 ± 4.2 |
| **Height (m)** |  |  |
| 1.75 ± 0.09 | 1.77 ± 0.09 | 1.78 ± 0.10 |
| **Mass (kg)** |  |  |
| 79.1 ± 19.9 | 84.2 ± 19.0 | 80.5 ± 16.4 |
| **Age At Surgery (y)** |  |  |
| 17.8 ± 2.3 | 19.5 ± 2.4 | 18.9 ± 2.9 |

**Table 2.**
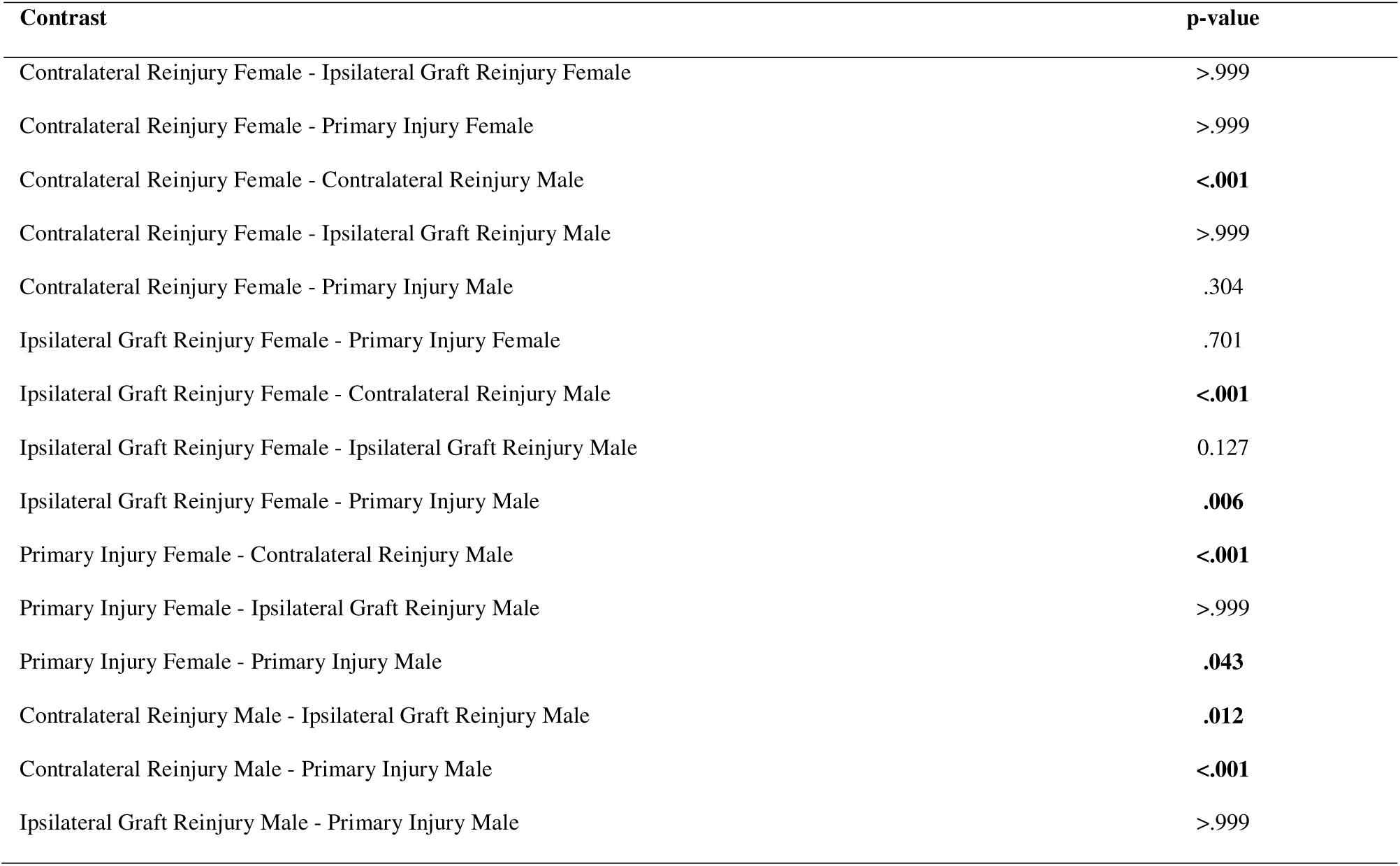
Post-hoc tests for the biological sex by injury status interaction.

## DISCUSSION

Psychological readiness following ACL injury plays a critical role in outcomes yet differences in the trajectory of ACL-RSI scores post-ACLR between patients following primary ACL injury and ACL reinjury remains unclear. This study aimed to compare ACL-RSI scores over time post-ACLR in male and female athletes aged 15-25 following primary ACL injury and ACL reinjury, hypothesizing that patients following ACL reinjury will have lower ACL-RSI scores, with females exhibiting lower scores than males. The main findings of this study highlight differences in psychological readiness based on injury status, biological sex, and time post-ACLR. Males consistently report higher ACL-RSI scores than females, which supports our hypothesis and agrees with the literature. Although we observed that patients with ipsilateral ACL graft reinjury exhibit lower ACL-RSI scores than those with contralateral ACL reinjury, patients did not exhibit lower ACL-RSI scores following ACL reinjury compared to primary ACL injury, which is counter to our hypothesis.

Evaluating the psychological response to ACL injury is critical. Patients exhibiting maladaptive psychological responses to ACL injury are less likely to adhere to rehabilitation programs, potentially manifesting as a lack of psychological readiness which is associated with a lower likelihood of RTS or return to pre-injury level of participation.(Ashton et al., 2020; Burland et al., 2019; Faleide & Inderhaug, 2023; Hsu et al., 2017; Webster & Feller, 2022; Webster et al., 2018; Xiao et al., 2023) Further, patients who exhibit worse psychological readiness to RTS may be at a higher risk for ACL reinjury (McPherson et al., 2019a; Paterno et al., 2018); although, patients with high psychological readiness to RTS may also be at an increased risk for reinjury due to a ‘cavalier’ mismatch in risk appraisal and earlier RTS post-surgery.(Zarzycki et al., 2024) Further, the pattern in psychological recovery post-ACLR is crucial and highlights the need for individualized monitoring throughout rehabilitation, as psychological readiness is dynamic and not inherently positive or negative; for example, patients following primary ACL injury who exhibit a constant or decreasing ACL-RSI score over time post-ACLR may also be at an increased risk for reinjury compared to patients who demonstrate an increase in score.(McPherson et al., 2019b; Zarzycki et al., 2024)

Generally, males tend to report higher ACL-RSI scores than females; although, the magnitude of the difference can vary and not all studies report significant sex-based disparities in ACL-RSI scores at all points in time post-ACLR.(Barth et al., 2023; Milewski et al., 2023; Obradovic et al., 2024; Sell et al., 2024) The results of our study support this and there are several suggested reasons for this sex-based difference. Males may be more likely to be motivated by the competitive nature of sports, and therefore may be more likely to exhibit and be comfortable with risk-taking behaviors associated with sports participation.(Bruder et al., 2023) Additionally, males report greater knee function, physical activity levels, and RTS rates post-ACLR in some studies, which could indirectly lead to or be associated with higher ACL-RSI scores.(Barth et al., 2023; Obradovic et al., 2024) Finally, it is possible that males might be less willing to honestly report maladaptive psychological responses, including fear and anxiety related to RTS, compared to females.(Milewski et al., 2023; Webster et al., 2018)

Duncan et al. (2023) demonstrated that patients who underwent revision ACLR had lower ACL-RSI scores compared those who had primary ACLR despite no other objective performance or self-reported functional differences between the two groups. This implies that a history of failed ACLR and the need for a revision inherently impacts a patient’s psychological response to injury. This difference may be influenced by several factors, including an increased fear of reinjury—specifically the fear of a third ACL injury—given their previous experience of reinjury, a decline in optimism about their second recovery, negative comparisons between their first and current rehabilitation, a further decrease in confidence, the mental and physical challenge of undergoing rehabilitation again and its effect on motivation, and unresolved maladaptive responses to their initial injury, all of which further disrupt their athletic identity due to the additional time lost.(Duncan et al., 2023)

Despite these logical hypotheses, we did not observe something comparable to Duncan et al. (2023). This may be due to several reasons, including Duncan et al. (2023) had a mean age of nearly 30 years old, which is much older than the high-risk population typically seeking a return to cutting and pivoting sports and older than the sample used in the present study. Older patients exhibit distinct psychological responses, including lower ACL-RSI scores, and RTS rates compared to younger patients.(Ashton et al., 2020; Faleide et al., 2021; McPherson et al., 2019a; Milewski et al., 2023; Webster et al., 2018; Zink et al., 2024; Zwolski et al., 2023) Additionally, Duncan et al. (2023) assessed ACL-RSI only at RTS, rather than its trajectory over time post-ACLR, and it is possible that ACL-RSI scores between the primary ACLR and reinjury patients converged and plateaued by this point post-ACLR, masking differences that occurred earlier in rehabilitation. In relation to this, monitoring the change in psychological readiness over time post-ACLR, as opposed to a single time point, has been suggested to be important, as the rate of change has been associated with a higher risk of ACL reinjury in younger patients.(McPherson et al., 2019b)

Assessing the trajectory of ACL-RSI scores of time post-ACLR could help identify patients with maladaptive psychological responses to injury at different stages of rehabilitation providing the opportunity to intervene before the time of RTS.(Barth et al., 2023) Therefore, because some of the patients included in the present study had ACL-RSI assessed at multiple points in time post-ACLR, those with lower ACL-RSI scores early on may have pursued additional support or interventions aimed at improving their psychological readiness to RTS leading to higher scores later during the course of rehabilitation; although, if this was done, this would have been done outside of our sports physical therapy clinics and was beyond our control. We also included primary ACLR patients who may or may not have RTS. If we had excluded primary ACLR patients who did not RTS, we may have observed higher ACL-RSI scores in this group given the relation between psychological readiness and probability of RTS. Finally, although it is logical to suspect lower ACL-RSI scores following ACL reinjury as demonstrated by Duncan et al. (2023), it is also possible that the second ACL injury could cause a change in a patient’s desire and motivation to RTS. For example, a patient could no longer desire to RTS, in particular at the same level, following the second ACL injury, in turn changing the context of psychological readiness to RTS. Therefore, careful consideration of RTS desire and motivation in future studies assessing this phenomenon is needed.

The lower ACL-RSI scores for ipsilateral ACL graft reinjury patients compared to contralateral ACL reinjury patients in the present study was not necessarily expected. In fact, in support of the opposite scenario, patient’s who sustain a contralateral ACL reinjury could have a fundamental shift in their perception of their body and susceptibility to injury, potentially impacting confidence and self-efficacy beyond just the previously injured knee.

Specifically, they may question their inherent anatomy and biomechanics, the effectiveness of their training and prevention programs, or the ability of their providers or coaches to identify and mitigate risk factors. Conversely, individuals who experience an ipsilateral ACL graft reinjury may simply attribute it to a failed surgery and nothing inherent to them. Therefore, future studies aiming to investigate ACL-RSI scores after ACL reinjury should carefully consider these perceptions.

There are some limitations of this retrospective study. This study analyzed data collected during routine clinical practice, which presents several challenges. The data were irregularly spaced as patients had a varying number of observations at different intervals.

Specifically, observations were more frequent between 4 to 8 months post-ACLR, resulting in a non-uniform distribution over time and sparse data, with most patients having two or fewer observations. Although our statistical approach is resilient to these issues, a more optimal approach would have involved standardized collection of ACL-RSI at fixed intervals, such as at 3, 6, 9, and 12 months post-ACLR, to achieve a more even distribution and greater density of observations per patient. Additionally, the influence of concurrent pathologies treated during ACLR, particularly meniscal repairs which could necessitate longer periods of non-weight bearing, were not accounted for but could significantly impact psychological readiness. The study cohort was limited to ACLR patients aged 15 to 25 years and did not control for age as a covariate; therefore, these results may not be generalizable to all age groups, despite this age range being most susceptible to ACL injuries. Data collection ceased at one-year post-ACLR, hence longer-term recovery trends remain unexplored. Surgical and rehabilitation protocols were also not considered in the analysis, which may influence psychological readiness. Although, this lack of consideration could be considered a strength as our data reflects a diverse range of surgical and providers, potentially enhancing the generalizability of our findings.

In conclusion, psychological readiness following ACL injury varies by injury status, biological sex, and time post-ACLR. While males reported higher ACL-RSI scores, we did not observe lower scores in reinjured patients compared to primary injuries. However, ipsilateral ACL graft reinjury patients exhibited lower scores than those with contralateral reinjury, suggesting different psychological responses based on reinjury type. These findings highlight the need for individualized psychological monitoring throughout rehabilitation.

## Ethical Statement

The Sanford Health Institutional Review Board granted this study an exemption (Study ID: 2390).

## Data Availability

Data produced in the present study are available upon reasonable request to the authors.

